# Clinical performance characteristics of the Swift Normalase Amplicon Panel for sensitive recovery of SARS-CoV-2 genomes

**DOI:** 10.1101/2021.10.22.21265255

**Authors:** Lasata Shrestha, Michelle J. Lin, Hong Xie, Margaret G. Mills, Shah A. Mohamed Bakhash, Vinod P. Gaur, Robert J. Livingston, Jared Castor, Emily A. Bruce, Jason W. Botten, Meei-Li Huang, Keith R. Jerome, Alexander L. Greninger, Pavitra Roychoudhury

## Abstract

Amplicon-based sequencing methods have been central in characterizing the diversity, transmission and evolution of SARS-CoV-2, but need to be rigorously assessed for clinical utility. Here, we validated the Swift Biosciences’ SARS-CoV-2 Swift Normalase Amplicon Panels using remnant clinical specimens. High quality genomes meeting our established library and sequence quality criteria were recovered from positive specimens with a 95% limit of detection of ≥ 40.08 SARS-CoV-2 copies/PCR reaction. Breadth of genome recovery was evaluated across a range of Ct values (11.3 – 36.7, median 21.6). Out of 428 positive samples, 406 (94.9%) generated genomes with < 10% Ns, with a mean genome coverage of 13,545X ± SD 8,382X. No genomes were recovered from PCR-negative specimens (n = 30), or from specimens positive for non-SARS-CoV-2 respiratory viruses (n = 20). Compared to whole-genome shotgun metagenomic sequencing (n = 14) or Sanger sequencing for the spike gene (n = 11), pairwise identity between consensus sequences was 100% in all cases, with highly concordant allele frequencies (R^2^ = 0.99) between Swift and shotgun libraries. When samples from different clades were mixed at varying ratios, expected variants were detected even in 1:99 mixtures. When deployed as a clinical test, 268 tests were performed in the first 23 weeks with a median turnaround time of 11 days, ordered primarily for outbreak investigations and infection control.

## INTRODUCTION

Since the deposition of the first SARS-CoV-2 whole genome sequence (NC_045512.1) in January 2020, over 4 million SARS-CoV-2 genomes have been deposited to public data repositories, far exceeding any other human pathogen^1,2^. Such a feat has been made possible due to advances in next-generation sequencing technologies that enable near real-time genomic surveillance^3–9^. Viral whole-genome sequencing (WGS) of laboratory-confirmed SARS-CoV-2 isolates is frequently used in outbreak investigations, deployment of public health interventions, development of vaccines and therapeutics, and evaluating vaccine and antiviral effectiveness against emerging variants^4–6,10–15^. Specific SARS-CoV-2 variants have been associated with higher viral loads, lower vaccine effectiveness, and worse outcomes such as mortality^16–23^. Notably, B.1.1.7 has been associated with increased disease severity, prolonged hospitalization, and higher mortality risk^17,19–24^. Additionally, recent studies have shown poorer outcomes for patients infected with variants B.1.351 and P.1^23,25^. In a clinical setting, identification of specific viral mutations can aid in the selection of monoclonal therapies^26–29^, and viral sequencing can be used to monitor the accumulation of mutations as a result of long-term viral replication in immunocompromised individuals^30^. As treatment regimens expand, validated WGS assays are needed not only for genomic surveillance but also for high-quality, clinically actionable data with rapid turnaround times.

Multiplexed amplicon sequencing methods have proven to be faster, more sensitive, and more cost-effective than shotgun and capture-based approaches, enabling genome recovery across a wide range of viral loads^31,32^. We previously tested one such panel from Swift Biosciences and demonstrated genome recovery up to a Ct of 36 across a broad range of isolates^33^. Designed against the SARS-CoV-2 Wuhan-Hu-1 complete genome (NC_045512.2), the Swift SNAP primer set amplifies 345 amplicons ranging from 116-255 bp (average 150 bp) in a single tube to cover the ∼30 kb SARS-CoV-2 genome. This assay can generate libraries in less than three hours using an input concentration of only 10 to 100+ viral copies for single-strand complementary DNA or double-strand complementary DNA synthesis. This can be followed by either manual normalization or Swift Biosciences’ proprietary enzymatic normalization of multiplexed libraries for equimolar pools.

For clinical use, sequencing assays need to be rigorously validated, documented, and performed in Clinical Laboratory Improvement Amendments (CLIA) accredited laboratories. However, no specific guidelines currently exist for the development and validation of WGS assays for SARS-CoV-2. Here we used existing FDA guidelines for NGS-based assays for germline diseases and development of antiviral products to clinically validate Swift Biosciences’ one-tube SARS-CoV-2 Swift Normalase Amplicon Panel (SNAP) Version 2.0–the first clinical validation of an NGS-based assay for WGS of SARS-CoV-2 to our knowledge^34,35^. Using clinical specimens, we evaluated the analytical sensitivity, analytical specificity, the limit of detection (LOD), accuracy, and precision (reproducibility and repeatability) of the assay, establishing acceptance criteria for sequencing libraries and output genomes. This assay is now available as a clinically orderable test with results returned to physicians to aid in disease management and treatment. It has been used in multiple vaccine trials, research studies, validation of other NGS-based assays, and in sequencing SARS-CoV-2 for public health surveillance and outbreak investigation^6,10,36–40^.

## MATERIALS AND METHODS

### Performance evaluation

We used existing FDA guidelines for designing, developing, and establishing the analytical validity of NGS-based tests for the diagnosis of suspected germline diseases and development of antivirals to clinically validate a multiplex amplicon sequencing method for WGS of SARS-CoV-2 (summarized in Supplementary Figure 1)^34^. We aimed to determine analytical sensitivity (limit of detection), analytical specificity, accuracy, and precision (reproducibility and repeatability) of the assay using remnant clinical specimens described below.

### Clinical specimens

Use of deidentified remnant clinical specimens from University of Washington Virology Laboratory (UWVL) for SARS-CoV-2 testing was approved by the University of Washington Institutional Review Board. Specimens were tested for SARS-CoV-2 using one of the following PCR assays: CDC-based Laboratory Developed Assays (LDA), Abbott m2000 (Abbott Laboratories, Chicago, IL, USA), Roche Cobas (Roche, Basel, Switzerland), or Hologic Panther/Panther Fusion (Hologic, Marlborough, MA, USA)^41,42^. Samples used for analysis of specificity were tested using UWVL respiratory virus panel to identify non-SARS-CoV-2 respiratory pathogens including respiratory syncytial virus (RSV), influenza virus type A (FluA), parainfluenza virus types 1, 2, and 3 (PIV1, PIV2, and PIV3), and human metapneumovirus (MPV)^43^.

Laboratory-confirmed specimens used in this study came from nasal or nasopharyngeal swabs collected in either PBS or viral transport media that had >500 μL volume remaining. This included SARS-CoV-2 positive specimens (*n* = 428), specimens negative for both SARS-CoV-2 and other respiratory viruses (*n* = 30), and specimens positive only for other respiratory viruses (*n* = 20). Water was used as a negative control and previously confirmed and sequenced SARS-CoV-2 positive specimens were used as positive controls.

### SARS-CoV-2 virus culture isolates

For detection of within-sample variation, we used two previously sequenced culture isolates from different clades: WA-UW-20236TM, EPI_ISL_4926371, Nextstrain clade 20A and WA-UW-19433TM, EPI_ISL_4926374, 20B. Viruses were isolated from original clinical specimens at the University of Vermont (UVM) BSL-3 facility under an approved Institutional Biosafety protocol as previously described^44^. Vero E6-TMPRSS2 cells (obtained from the JCRB Cell Bank, JCRB No. JCRB1819) were inoculated with 100 μL of clinical sample, inoculated for 1 hour at 37ºC with rocking, washed with PBS, and overlaid with 1 ml standard Vero media containing 2% FBS. Wells were monitored daily for the appearance of cytopathic effect (CPE) and successful SARS-CoV-2 isolation was confirmed by immune-focus assay^45^. Once CPE was observed, supernatants were clarified to remove cellular debris and prepared for RNA extraction by mixing 1:1 with Qiagen Buffer AVL. The high viral loads in supernatants post viral isolation (Ct 10 and 9.5 respectively) allowed preparation of multiple dilutions and mixtures. The consensus sequences for the two samples differed at a total of 16 positions, with 6 mutations unique to the 20A specimen (C4633T, C10965T, T14643C, A20268G, C22482T, C28854T), 10 mutations unique to the 20B specimen (G2144T, G3824A, G13348T, C15933T, G16968T, T19839C, G28083T, GGG(28881-28883)AAC), and 4 mutations (C241T, C3037T, C14408T, A23403G) were common to both samples. All positions are relative to the SARS-CoV-2 isolate Wuhan-Hu-1 reference sequence (NC_045512.2).

### RNA Extraction

RNA was extracted from 200 μL of each specimen (nasal or nasopharyngeal swabs in PBS or VTM) and eluted to 100 μL on the MagNAPure LC (Roche, Basel, Switzerland) using the manufacturer’s instructions. This was followed by a confirmatory RT-PCR using AgPath-ID One-Step RT-PCR kit (Life Technologies, Carlsbad, CA, USA) with CDC primers and probes for N1/N2 gene following protocol and concentrations defined in Corman et al^41,46^. Specimens with N1/N2 cycle threshold (Ct) ≤ 18 were diluted to Ct 22-24 to prevent inhibition in downstream procedures.

### Swift SNAP library preparation and quality control

Using SuperScript™ IV First-Strand Synthesis System (ThermoFisher, Waltham, MA, USA), 11 μL of extracted RNA was subjected to single-strand complementary DNA (sscDNA) synthesis, and 10 μL of the resulting sscDNA was used for library preparation using the Swift SARS-CoV-2 SNAP Version 2.0 kit (Swift Biosciences™, Ann Arbor, MI, USA). First strand synthesis and library preparation protocols were automated on liquid handlers – Sciclone® G3 NGSx iQ™ Workstation (PerkinElmer™, Waltham, MA, USA) following manufacturer’s instructions (rev 9-1, https://www.protocols.io/view/uw-virology-swift-snapv2-protocol-byw4pxgw). Libraries were quantified on a VICTOR Nivo Multimode Microplate Reader (PerkinElmer, Waltham, MA, USA) using the Quant-iT dsDNA High Sensitivity kit (Life Technologies, Carlsbad, CA, USA). Library quality was inspected on the Agilent 4200 TapeStation (Agilent Technologies, Santa Clara, CA, USA) using TapeStation D1000 DNA ScreenTape. Libraries with concentrations ≥ 1.35 ng/μL on the plate reader and ≥ 0.5 ng/μL (region concentration from 250 bp to 750 bp) on the TapeStation were deemed accepted for sequencing. These cutoffs were determined based on multiple runs of negative controls and expected ranges of amplicon sizes. Libraries satisfying these criteria were enzymatically normalized using Normalase to generate equimolar pools using the Swift SARS-CoV-2 SNAP Version 2.0 kit. Generated pools were sequenced on the Illumina NextSeq 500 (Illumina, San Diego, CA, USA) using the NextSeq 500/550 High Output Kit v2.5 (2×150 Cycles). Positive and negative controls were included on each run, and only sequencing runs with > 50% reads passing filter (PF) and > 60% of bases exceeding Phred quality scores of 30 (Q30) were accepted.

### Instrument validation

After initial validation on the Illumina NextSeq 500, we ran parallel sets of libraries on Illumina NextSeq 2000 and Illumina MiSeq instruments using 2×150 kits. Run, library, and genome quality metrics were assessed to ensure they meet passing criteria and consensus genomes and allele frequencies were compared across the two runs.

### Metagenomic sequencing

Shotgun metagenomic sequencing (mNGS) was performed as previously described^47,48^. Briefly, extracted RNA from selected positive specimens (Ct ≤ 25) subjected to double-stranded cDNA synthesis using the SuperScript™ IV First-Strand Synthesis System followed by second-strand synthesis using Sequenase v2.0 (ThermoFisher, Waltham, MA, USA) and AMPure XP (Beckman Coulter Life Sciences, Brea, CA, USA) bead-based purification. Libraries were prepared using Nextera Flex (Illumina) following manufacturer’s protocols and sequenced on the NextSeq 500/550 (Illumina) using the High Output Kit v2.5 (75 cycles).

### Sanger sequencing (spike gene)

Three amplicons covering the Spike coding region (NC_045512.2: 21252-22861, 22559-24395, 24072-25572) were generated for each sample using SuperScript III One-Step PCR Mix (Supplementary material). Amplicons were purified using Qiagen columns, eluted with 50μl of elution buffer, and 2μl of each purified sample was run on a FlashGel (Lonza). Samples with visible bands were Sanger sequenced using 4 to 8 separate sequencing primers (IDT) and compared against corresponding NGS samples.

### Genome assembly and data analysis

Genomes from Swift SNAP libraries were assembled using a custom pipeline, TAYLOR (https://github.com/greninger-lab/covid_swift_pipeline) described previously^33,39^. Briefly, raw reads were adapter- and quality-trimmed using BBDuk (https://jgi.doe.gov/data-and-tools/bbtools/), aligned to the Wuhan-Hu-1 reference genome (NC_045512.2), and trimmed of PCR primers using Primerclip (https://github.com/swiftbiosciences/primerclip). Consensus genomes and variants were called using BCFtools^49^. For a genome to pass acceptability criteria we required: > 1 million raw reads, >750x mean genome coverage, >1000x mean spike gene coverage, 100% of spike gene with at least 200x coverage, and <10% unknown bases (Ns) in the final consensus sequence. A minimum of 6x per base coverage was required to call a non-N base in the consensus sequence.

For Sanger sequencing, reads were imported into Geneious (v9.1.8, Biomatters, Auckland, New Zealand), trimmed with an error probability limit of 0.005, mapped to the reference Spike sequence (NC_045512.2: 21563-25384), and manually trimmed to the start and stop codons for Spike. Trimmed reads were *de novo* assembled and the final Sanger consensus sequence for Spike was extracted. Samples that passed QC for Swift and had a complete spike consensus sequence by Sanger were included in the comparison. Consensus sequences for spike were aligned using MAFFT v7.45, and the number of pairwise nucleotide differences between sequences compared.

Shotgun metagenomic sequencing libraries were also analyzed using the same bioinformatic pipeline (TAYLOR) but with settings adjusted to omit primer trimming. Consensus sequences were aligned using MAFFT v7.45 and pairwise identity computed by comparing nucleotide differences between Swift and shotgun sequences. Allele frequencies for all snps called by the Taylor pipeline at 1% or higher were compared between Swift and shotgun samples.

Probit analysis to determine limit of detection calculations was performed in SPSS (v26, IBM, Armonk, NY). Other statistical analyses and data visualizations were performed using R^50^.

### Clinical test result reporting

For clinical orders (https://testguide.labmed.uw.edu/public/view/SARSEQ and https://testguide.labmed.uw.edu/public/view/SAREPI), we perform downstream analysis on consensus sequences for samples that pass all QC criteria described above. Consensus sequences are analyzed using Pangolin (https://pangolin.cog-uk.io/) to obtain PANGO lineage designations, and Nextclade (https://clades.nextstrain.org/) to generate Nextstrain clade assignments and a list of mutations. The mutation list generated from Nextclade is parsed to extract amino acid changes in Spike. For SARSEQ orders, the PANGO lineage, Nextstrain clade, and list of amino acid changes in Spike is returned along with a free text interpretation of these results. The interpretation includes information about variants of concern/variants of interest (VOC/VOI) and any known phenotypic effects associated with specific mutations such as increased transmissibility, or reduced neutralization by monoclonal antibodies (mAbs). Example reports are shown in Supplementary Figure 3. For SAREPI orders (requested for outbreak/cluster investigations), consensus sequences passing QC criteria are aligned using MAFFT v7.45 and the total number of pairwise nucleotide differences between sequences is reported as a table along with SARSEQ results.

### Bioinformatic pipeline validation

To assess the performance of our bioinformatic pipeline and data processing steps, we used a set of benchmark datasets from the CDC (https://github.com/CDCgov/datasets-sars-cov-2). These datasets contain raw fastq files for samples generated using the ARTIC protocol (https://artic.network/ncov-2019), with results indicating PANGO lineages, and designation as VOC/VOI. We ran our TAYLOR pipeline described above on the benchmark datasets, with settings to perform primer trimming for ARTIC amplicon primers and compared consensus sequences and lineage designations against the results provided. This analysis is performed each time there is a major version change in our bioinformatic pipeline.

## RESULTS

### Swift SNAP assay validation using clinical specimens

We performed rigorous assay validation using deidentified clinical specimens received at the UWVL between 09/21/2020 and 10/19/2020. We defined run-level and sample-level acceptability criteria (see Methods) ensuring multiple quality control checks at each stage of library preparation and sequencing. All sequencing runs successfully met run quality metrics with 93.24% average %Q30 (SD = 4.46%), 92.95% average %PF (SD = 5.65%), no amplification of negative controls, and successful recovery of positive controls.

### Limit of detection of Swift SNAP assay is 40 copies per PCR reaction

To determine the limit of detection of the assay, serial dilutions of extracted RNA were prepared from a positive SARS-CoV-2 clinical sample (WA-UW-29702, EPI_ISL_603255). Multiple replicate libraries (n=4 to 20 replicates, Table 1) were prepared for each dilution with target concentrations ranging from 500 copies/reaction to 5 copies/reaction. The concentrations of the prepared dilutions were then verified by qRT-PCR and measured concentrations ranged from 698.0 to 7.8 copies/reaction. At concentrations of 82 copies/reaction and higher, high-quality genomes were recovered in 100% of attempted replicates (n = 4 to 20, Table 1). The lowest concentration at which acceptable genomes were recovered 95% of the time was defined as the limit of detection (LOD) and determined by Probit analysis to be 40.08 copies/reaction (Table 1, Supplementary Table 1).

**Table 1.** Limit of detection determination using serial dilutions of a positive specimen in replicates

| Concentration<br>(copies/reaction) | Number of<br>replicates | Passing genomes<sup>a</sup> | % Passing<sup>b</sup> |
| --- | --- | --- | --- |
| **698.0** | 4 | 4 | 100 |
| **309.1** | 20 | 20 | 100 |
| **157.5** | 20 | 20 | 100 |
| **82.0** | 20 | 20 | 100 |
| **51.1** | 20 | 19 | 95 |
| **14.8** | 5 | 4 | 80 |
| **7.9** | 5 | 2 | 40 |

<sup>a</sup>Number of genomes satisfying all acceptability criteria
<sup>b</sup>Percentage of genomes satisfying all acceptability criteria

### No cross-reactive amplification of non-SARS-CoV-2 respiratory virus genomes

We evaluated assay specificity by checking for cross-reactivity to non-SARS-CoV-2 respiratory viruses using clinical specimens positive for seasonal coronaviruses, rhinovirus, human parainfluenza viruses (HPIV-1, HPIV-2, HPIV-3, HPIV-4), respiratory syncytial virus (RSV), adenovirus, metapneumovirus, influenza A, and influenza B viruses (n = 20 samples total, Supplementary Table 2)^51^. Only 15/20 (75%) non-SARS-CoV-2 or other respiratory virus-positive specimens met the library QC criteria to be sequenced. No recoverable genomes were obtained from any of the 15 specimens and the number of mapped reads ranged between 0 and 3,114 reads which represented <1% of post-trimmed reads in all cases (Supplementary Table 2).

### Recovery of high-quality genomes from clinical samples across a wide range of viral loads

Breadth of genome recovery was evaluated using 428 SARS-CoV-2 positive clinical samples (nasopharyngeal, nasal, or oropharyngeal swabs) across a range of Ct values (11.3 – 36.7, median 21.6) and 30 SARS-CoV-2 negative clinical samples. Out of 428 positive samples, 406 (94.9%) generated genomes with < 10% Ns, with a mean genome coverage of 13,545 ± SD 8,382 (Supplementary Table 3).

Of the 30 samples that were PCR-negative for SARS-CoV-2, 17 did not meet quality control criteria for successfully amplification during library preparation and were not taken forward for sequencing. Of the remaining 13 sequenced samples, the number of mapped reads ranged between 0 and 5271, representing < 0.5% of post-trimmed reads. No genomes were recovered from any of these samples (Supplementary Table 4).

### Accurate detection of within-sample variation

RNA was extracted from two SARS-CoV-2 cultured viral isolates from distinct clades (WA-UW-20236TM, Nextstrain clade 20A and WA-UW-19433TM, 20B) with multiple distinguishing mutations (see Methods). Mixtures were prepared with ratios of 20A:20B ranging from 0:100 to 50:50 to 100:0. Mutations were called against the Wuhan-Hu1 reference sequence (NC_045512) and filtered to accept mutations with allele frequency > 1%. All expected mutations were identified at mixing ratios of 5:95 and above (Table 2, Figure 1). At mixing ratios of 1:99, 4 out of 6 of the expected low-frequency 20A mutations were identified. In the remaining two mixtures, variants were present but at allele frequencies lower than 1%, as would be expected based on mixing at 1:99 (mean af and depth: C4633T 0.78% 10025x; C10965T 0.96% 10028x, n = 3 replicates).

**Table 2.** Genome recovery from SARS-CoV-2 PCR-positive and PCR-negative clinical specimens

| **Ct** | **# Samples** | **Passing genomes** |
| --- | --- | --- |
| **<15** | 60 | 56 (93%) |
| **15-20** | 123 | 118 (96%) |
| **20-25** | 109 | 106 (97%) |
| **25-30** | 97 | 91 (94%) |
| **>30** | 39 | 35 (90%) |
| **Negative** | 30 | 0 |

**Figure 1.**
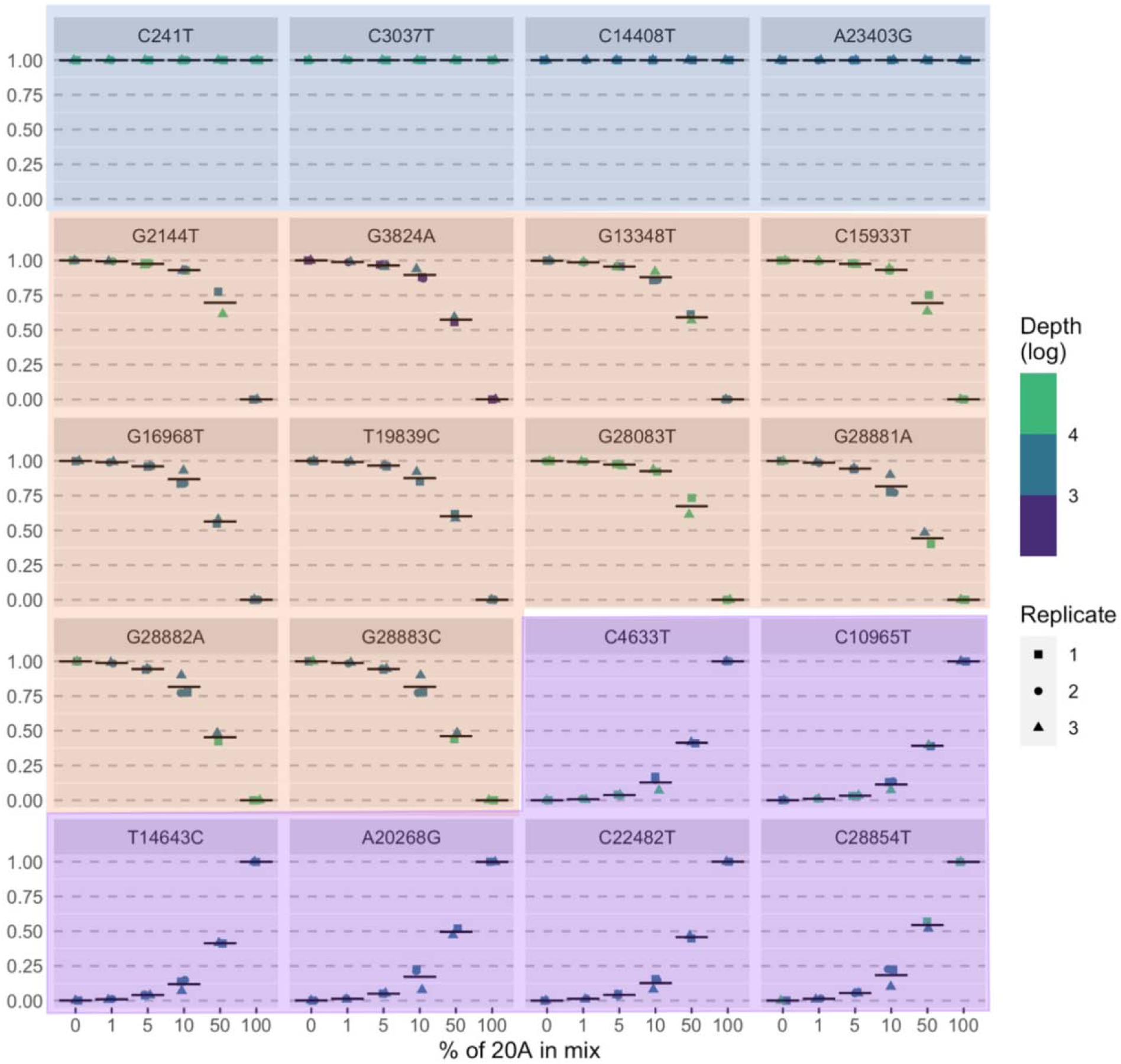
Measured allele frequency (y-axis) for all expected mutations (n =20) in sample mixtures described in Table 2. Blue panels = mutations common to 20A and 20B samples, orange panels = mutations expected in 20B sample, purple panel = mutations expected in 20A sample.

**Table. 2.** Variant detection in mixtures prepared from two samples: WA-UW-20236TM (20A) and WA-UW-19433TM (20B).

| **Mix** | **% 20A specimen** | **% 20B specimen** | **Common variants detected (4 total\*)** | **20A variants detected (6 total\*\*)** | **20B variants detected (10 total\*\*\*)** |
| --- | --- | --- | --- | --- | --- |
| **1** | 0 | 100 | 4 | 0 | 10 |
| **2** | 1 | 99 | 4 | 4 | 10 |
| **3** | 5 | 95 | 4 | 6 | 10 |
| **4** | 10 | 90 | 4 | 6 | 10 |
| **5** | 50 | 50 | 4 | 6 | 10 |
| **6** | 100 | 0 | 4 | 6 | 0 |

\*Expected variants common to both samples (n=4): C241T, C3037T, C14408T, A23403G
\*\*Variants in 20A sample (n=6): C4633T, C10965T, T14643C, A20268G, C22482T, C28854T
\*\*\*Variants in 20B sample (n=10): G2144T, G3824A, G13348T, C15933T, G16968T, T19839C, G28083T, GGG(28881-28883)AAC

### High concordance with Sanger and shotgun metagenomic sequencing

To test whether assay results agreed with alternative sequencing methodologies, a subset of clinical positive specimens sequenced by Swift was subjected to Sanger sequencing of the spike gene and shotgun metagenomic next-generation sequencing (mNGS). Full-length spike consensus sequences obtained by Sanger sequencing were compared against their corresponding Swift sequences for 14 positive specimens with Ct ≤ 30. All 14 specimens selected for Sanger sequencing passed genome acceptability criteria by Swift while 11/14 (78.6%) generated a complete Sanger sequence for spike (Supplementary Figure 2). Pairwise sequence identity between spike consensus sequences for Sanger and Swift was 100% for all 11 samples (Supplementary Table 5).

Shotgun mNGS was performed on 14 samples with Ct ≤ 25, and consensus sequences were obtained for all 14 samples with mean genome coverage 4,304x (SD=9,886x) and 0 – 1.3 %Ns (Supplementary Table 6). Pairwise sequence identity between shotgun and Swift sequences was 100% for all 14 sample pairs. For all single nucleotide variants identified at 1% or higher, allele frequencies were highly concordant between Swift and mNGS libraries (Figure 2A, R^2^ = 0.99, Pearson correlation coefficient = 0.99, p < 2.2 e-16). Of these variants, only 2 out of 7421 (0.03%) had allele frequencies differing more than 20% between library preparation methods, attributable to poor subgenomic RNA mapping in the Swift library or noisy ends of reads in the metagenomic library of one sample (WA-UW-29702). Across the genome, depth of coverage ranged from 0 to 370,627x for Swift and 0 to 130,281x for mNGS (Figure 2B), with Pielou’s evenness > 0.99 for both mNGS and Swift libraries. The mean whole-genome coverage in the Swift libraries (15,631x) was greater than in the mNGS libraries (9,981x). Notably, in one sample (WA-UW-29751), we observed a drop in coverage at positions 27823-28233 in both Swift and mNGS samples corresponding to a 411-nucleotide deletion in ORF7b/8 that has been found in other samples from WA (data not shown).

**Figure 2.**
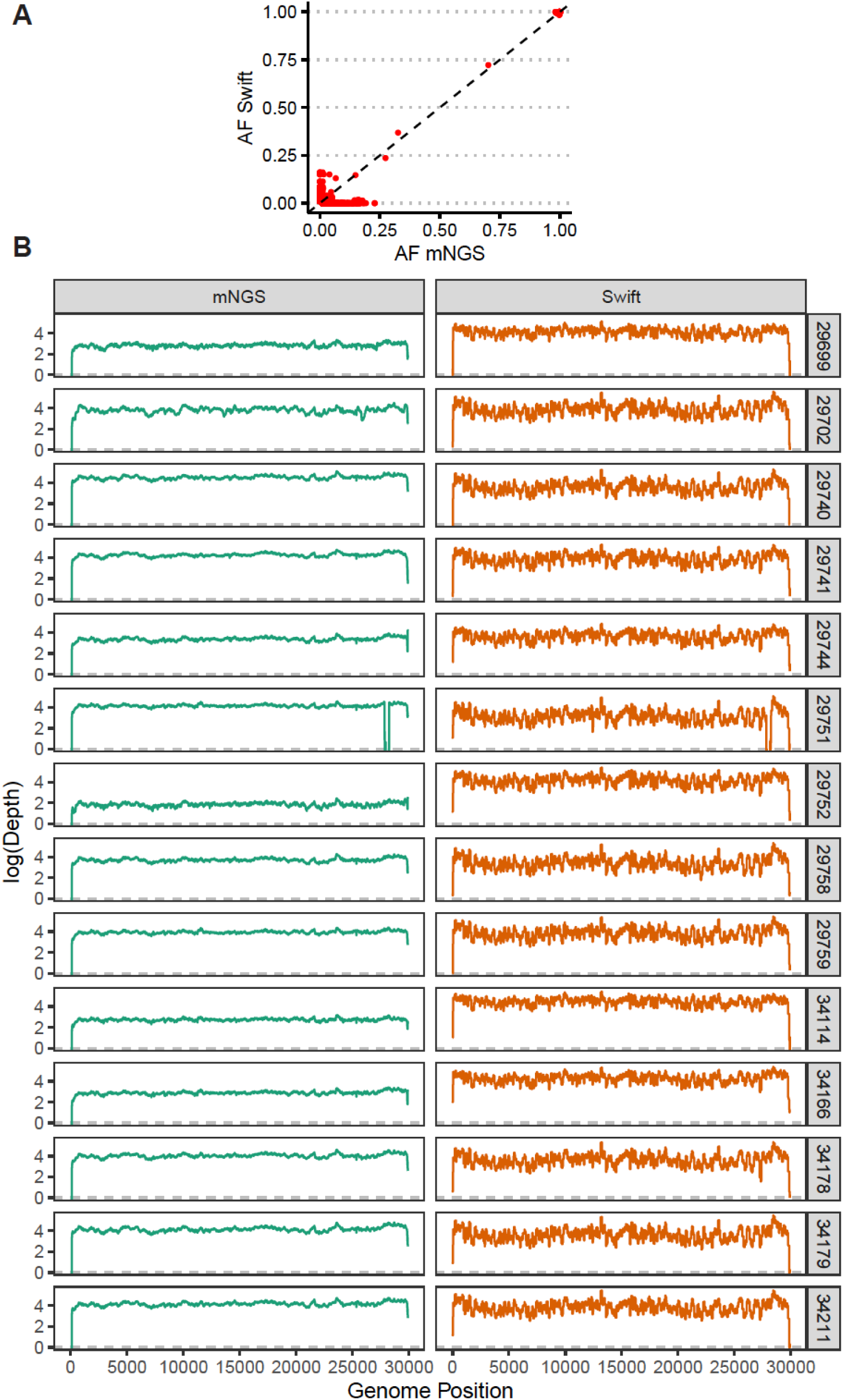
Swift vs shotgun mNGS. A). Allele frequencies were highly concordant between Swift and mNGS libraries (n = 14 samples); B) Comparing depth of coverage (log_10_ reads) vs. genomic position relative to NC_045512 for specimens sequenced with both SWIFT and mNGS.

### High assay precision (repeatability and reproducibility)

Precision was evaluated by testing the concordance of assay results and quality metrics for specimens tested multiple times on the same run (repeatability) and on different runs (reproducibility). For repeatability, two replicate libraries were prepared for 10 SARS-CoV-2 positive specimens on the same library preparation plate on the liquid handler and the same sequencing run. For reproducibility, 10 SARS-CoV-2 positive specimens had libraries prepared three days later and sequenced on a separate run by a different technician. All the replicates of the specimen both on the same run and on different runs yielded acceptable genomes with highly correlated allele frequencies as shown in Figure 3. Assay results were also highly reproducible across different instruments including Illumina NextSeq 500, 2 separate NextSeq 2000s, and Miseq (Supplementary Figures 4, 5, 6).

**Figure 3.**
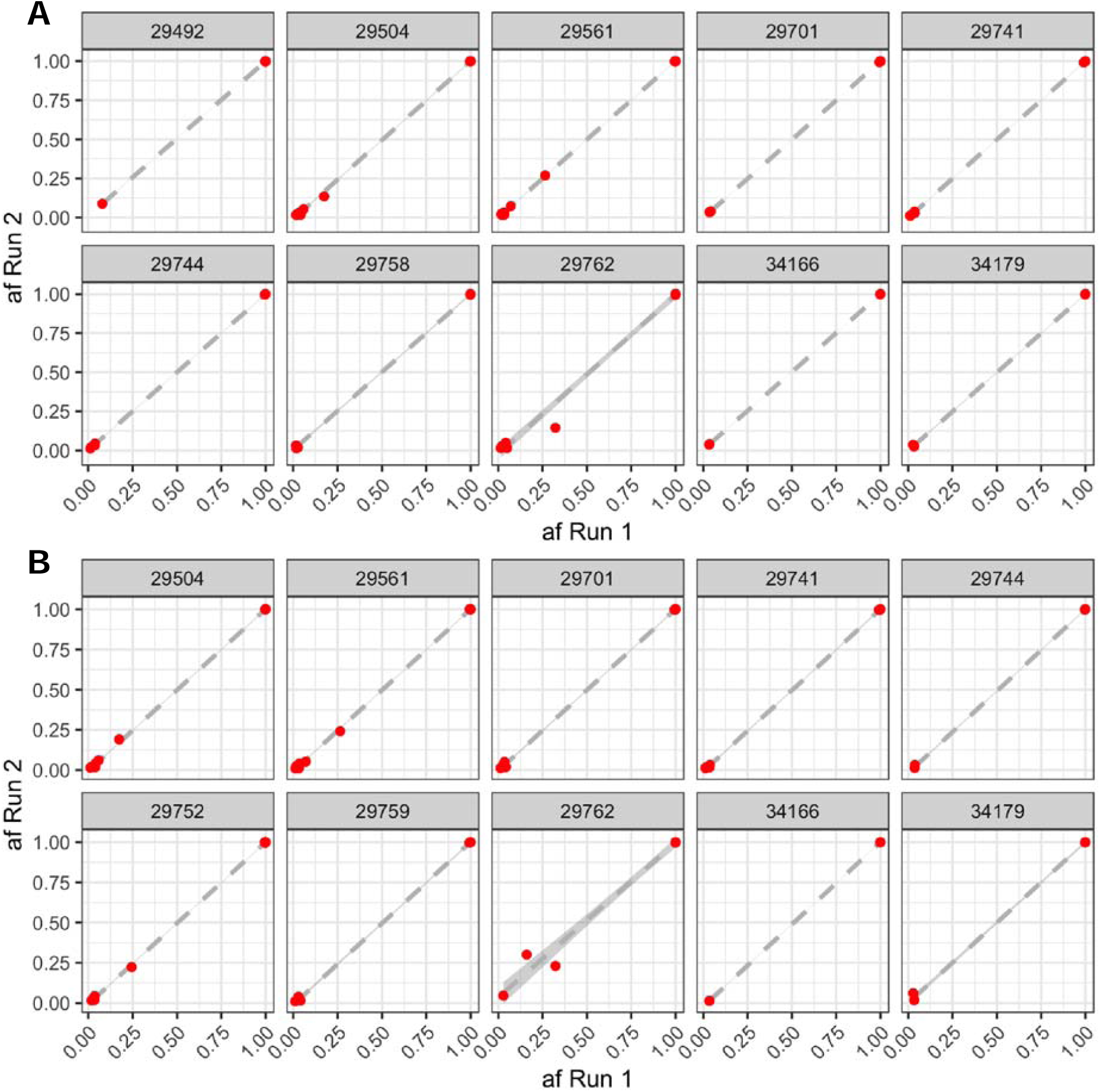
Highly reproducible allele frequencies across replicate libraries of 12 SARS-CoV-2 positive specimens sequenced on the same run (A), and on different runs on a different day by a different technician. Dashed grey line shows line of best fit by linear regression.

### Accurate identification of VOC and non-VOC lineages with benchmark datasets

We evaluated the performance of our data processing pipeline using CDC benchmark datasets of samples confirmed to be VOC/VOI (n = 16 and non-VOC/VOI (n = 39) sequences. The number of raw fastq reads ranged from 41,436 to 6,798,138 (median 602,508) across both datasets (Supplementary Table 7). After processing through our bioinformatic pipeline, consensus genomes with < 10% Ns were obtained for all samples, with mean genome coverage between 159x - 28,122x (median 2,598x), and mean spike gene coverage between 138x - 22,099x (median 1,954x). Pango and Nextclade lineages were compared for consensus sequences generated by our pipeline against benchmark deposited consensus genomes. Lineages agreed in 16/16 VOC/VOI samples and 37/39 non-VOC/VOI samples. In the two non-VOC samples where the lineages did not agree, the discrepancy was found to be due to mixed populations being reported using IUPAC ambiguities in the benchmark set vs. the majority consensus base being reported by TAYLOR. After masking ambiguous sites, consensus sequences in these discrepant samples showed 100% pairwise identity.

### Deployment of a high-throughput NGS assay in a clinical setting: the UW Virology experience

The UW Virology SARS-CoV-2 clinical sequencing assay went live on March 3, 2021 and in the first 23 weeks since launch, a total of 268 cases were processed (Figure 4). The median turnaround time, defined as order to first result, was 11 days. Tests were indicated for infection control (infections among patient-facing staff members), confirmation of patient-staff transmission, and cluster investigations. The diagnostic yield of Variants of Concern (VOC) identified in this period was 64% (171 samples). Among clinical orders, the first sample of B.1.617.2/Delta was detected in week 12.

**Figure 4.**
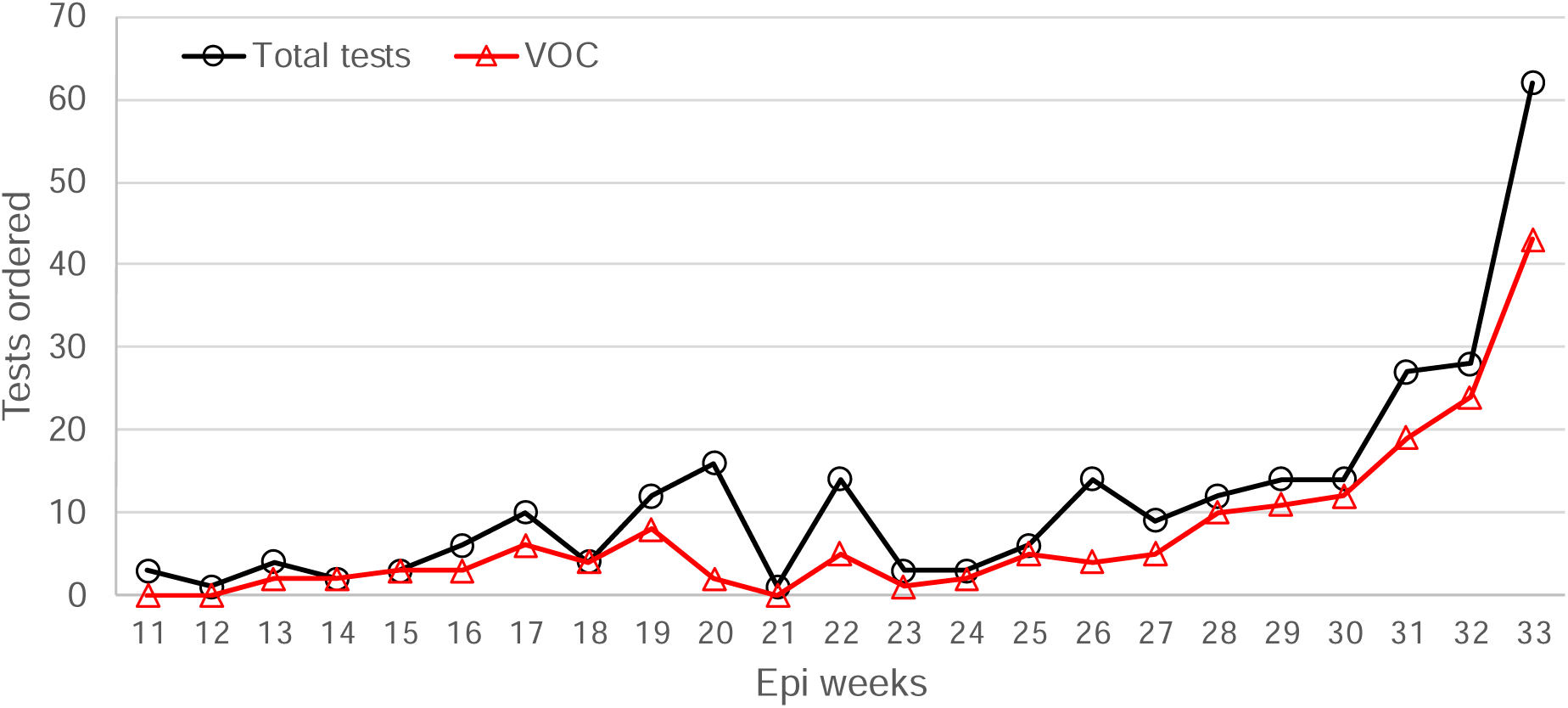
Weekly SARS-CoV-2 clinical whole genome sequencing test volumes, along with the number of VOCs detected in each week’s batch.

Semi-annual proficiency testing was performed via a sample exchange with a peer CLIA/CAP-certified laboratory in which six blinded specimens were exchanged and sequenced. Unblinding was performed following the completion of the study to compare the strains identified and the Spike gene mutations detected. Among proficiency testing samples exchanged, 100% of PANGO lineages and 98% of Spike gene mutations were identified correctly.

## DISCUSSION

Tiling multiplex PCR amplicon panels are an attractive option for high-throughput sequencing of SARS-CoV-2 as they allow recovery of genomes from a wide range of viral loads with relatively short workflows. Here we performed systematic validation of the Swift SNAP protocol using FDA guidelines for other NGS-based assays, in order to deploy the assay for routine clinical use in a CLIA and CAP-approved facility. The Swift SNAP assay demonstrated high sensitivity with recovery of full-length genomes at viral loads as low as 40.08 copies per reaction, with the ability to faithfully recover allele frequencies in mixed populations. The latter feature is of particular importance in quantifying within-host variation to track mutations with potential clinical significance following vaccination or treatment. Recent studies have suggested that amplicon sequencing provides a more accurate representation of minor allele frequencies than capture-based sequencing^31,52^.

The Swift SNAP panel produces short amplicons under 255bp that can be directly sequenced with short-read technologies like Illumina without additional fragmentation. In contrast, the popular low-cost ARTIC panel generates amplicons of size ∼400bp that require fragmentation prior to sequencing on Illumina platforms, or direct sequencing via Oxford Nanopore Technologies (ONT)^53,54^. In addition, the larger number of primers and overlapping amplicons in SNAP results in fewer amplicon dropouts compared to ARTIC-based methods^54^.

Given the wide range of viral loads among SARS-CoV-2 specimens, it can be challenging to optimize multiplex amplicon panels to simultaneously achieve high sensitivity and evenness of genome coverage. We found the manufacturer recommended settings to be adequate for genome recovery across the range of viral loads we see in a majority of clinical specimens. At very low viral loads, the high background of non-viral RNA can make genome recovery challenging. In addition, the Swift SNAP protocol includes an optional proprietary enzymatic normalization (Normalase) step, which results in equimolar pools with even coverage across samples and amplicons. This allows for batching of samples with minimal sample reorganization. While additional cycles of amplification may help stretch analytical sensitivity, this can result in an accumulation of primer dimers that can affect overall run quality and may amplify trace contaminants^54^.

One of the limitations of short-read sequencing using reference-based approaches is the reduced ability to identify large structural variants, long indels, and tandem duplicates. Long deletions often manifest as low coverage regions masked with ambiguous bases (Ns) and require manual review to confirm that they are deletions vs. amplicon dropouts. In the current study, we saw one example of this, and we have previously identified similar deletions in ORFs 7a, 7b, and 8 using Swift^55^.

Taken together, our results demonstrate the high sensitivity, specificity, and reproducibility of the Swift SNAP amplicon panel for SARS-CoV-2, which make it ideal for clinical applications. Our protocol is available at https://www.protocols.io/view/uw-virology-swift-snapv2-protocol-byw4pxgw with options for automation via robotic liquid handling systems. In addition, our study provides a framework for validating amplicon sequencing methods which have proved to be an important tool in our fight against COVID-19 and will be for other emerging pathogens.

## Data Availability

All data produced in the present work are contained in the manuscript and sequences have been deposited to GISAID.

## Acknowledgements

This work was supported by NIH AI027757 to P.R. and NIH P30GM118228-04 to E.A.B. Computational analyses were supported by Fred Hutch Scientific Computing (NIH ORIP grant S10OD028685) and UW Laboratory Medicine Informatics. We are grateful to Noah Hoffman and Niklas Krumm for guidance on cloud-based storage and analysis, to the CDC TOAST team for support with the CDC benchmark datasets, and Kanika Sharma (PerkinElmer) for automation help.

## Notes

Competing interests: ALG reports contract testing from Abbott Laboratories and research support from Gilead and Merck, outside of the described work. All other authors report no other financial or non-financial competing interests for this work.

### Competing Interest Statement

ALG reports contract testing from Abbott Laboratories and research support from Gilead and Merck, outside of the described work. All other authors report no other financial or non-financial competing interests for this work.

### Author Declarations

The Institutional Review Board of the University of Washington approved this work and provided a waiver of informed consent for the use of deidentified patient specimens in this work.

## References

1. Elbe, S. & Buckland-Merrett, G. Data, disease and diplomacy: GISAID’s innovative contribution to global health. Global challenges (Hoboken, NJ) 1, 33–46 (2017).

2. Moustafa, A. M. & Planet, P. J. Jumping a Moving Train: SARS-CoV-2 Evolution in Real Time. Journal of the Pediatric Infectious Diseases Society (2021) doi:10.1093/jpids/piab051.

3. Paden, C. R. et al. Rapid, Sensitive, Full-Genome Sequencing of Severe Acute Respiratory Syndrome Coronavirus 2. Emerging infectious diseases 26, 2401–2405 (2020).

4. Fauver, J. R. et al. Coast-to-Coast Spread of SARS-CoV-2 during the Early Epidemic in the United States. Cell 181, 990-996.e5 (2020).

5. Deng, X. et al. Genomic surveillance reveals multiple introductions of SARS-CoV-2 into Northern California. Science 369, 582 LP –587 (2020).

6. Bedford, T. et al. Cryptic transmission of SARS-CoV-2 in Washington state. Science 370, 571 LP –575 (2020).

7. Korber, B. et al. Tracking Changes in SARS-CoV-2 Spike: Evidence that D614G Increases Infectivity of the COVID-19 Virus. Cell 182, 812-827.e19 (2020).

8. Tang, J. W., Toovey, O. T. R., Harvey, K. N. & Hui, D. D. S. Introduction of the South African SARS-CoV-2 variant 501Y.V2 into the UK. The Journal of infection S0163-4453(21)00030-X (2021) doi:10.1016/j.jinf.2021.01.007.

9. Washington, N. L. et al. Genomic epidemiology identifies emergence and rapid transmission of SARS-CoV-2 B.1.1.7 in the United States. medRxiv 2021.02.06.21251159 (2021) doi:10.1101/2021.02.06.21251159.

10. Lieberman, N. A. P. et al. In vivo antiviral host transcriptional response to SARS-CoV-2 by viral load, sex, and age. PLOS Biology 18, e3000849 (2020).

11. Toyoshima, Y., Nemoto, K., Matsumoto, S., Nakamura, Y. & Kiyotani, K. SARS-CoV-2 genomic variations associated with mortality rate of COVID-19. Journal of Human Genetics 65, 1075–1082 (2020).

12. Lopez Bernal, J. et al. Effectiveness of Covid-19 Vaccines against the B.1.617.2 (Delta) Variant. New England Journal of Medicine (2021) doi:10.1056/NEJMoa2108891.

13. Madhi, S. A. et al. Efficacy of the ChAdOx1 nCoV-19 Covid-19 Vaccine against the B.1.351 Variant. New England Journal of Medicine 384, 1885–1898 (2021).

14. Chemaitelly, H. et al. mRNA-1273 COVID-19 vaccine effectiveness against the B.1.1.7 and B.1.351 variants and severe COVID-19 disease in Qatar. Nature Medicine (2021) doi:10.1038/s41591-021-01446-y.

15. Nasreen, S. et al. Effectiveness of COVID-19 vaccines against variants of concern in Ontario, Canada. medRxiv 2021.06.28.21259420 (2021) doi:10.1101/2021.06.28.21259420.

16. Frampton, D. et al. Genomic characteristics and clinical effect of the emergent SARS-CoV-2 B.1.1.7 lineage in London, UK: a whole-genome sequencing and hospital-based cohort study. The Lancet Infectious Diseases (2021) doi:https://doi.org/10.1016/S1473-3099(21)00170-5.

17. Fajnzylber, J. et al. SARS-CoV-2 viral load is associated with increased disease severity and mortality. Nature Communications 11, 5493 (2020).

18. Hung, I. F. N. et al. Viral loads in clinical specimens and SARS manifestations. Emerging infectious diseases 10, 1550–1557 (2004).

19. Bager, P. et al. Risk of hospitalisation associated with infection with SARS-CoV-2 lineage B.1.1.7 in Denmark: an observational cohort study. The Lancet Infectious Diseases (2021) doi:https://doi.org/10.1016/S1473-3099(21)00290-5.

20. Patone, M. et al. Mortality and critical care unit admission associated with the SARS-CoV-2 lineage B.1.1.7 in England: an observational cohort study. The Lancet Infectious Diseases (2021) doi:https://doi.org/10.1016/S1473-3099(21)00318-2.

21. Davies, N. G. et al. Increased mortality in community-tested cases of SARS-CoV-2 lineage B.1.1.7. Nature 593, 270–274 (2021).

22. Challen, R. et al. Risk of mortality in patients infected with SARS-CoV-2 variant of concern 202012/1: matched cohort study. BMJ 372, n579 (2021).

23. Paredes, M. I. et al. Associations between SARS-CoV-2 variants and risk of COVID-19 hospitalization among confirmed cases in Washington State: a retrospective cohort study. medRxiv 2021.09.29.21264272 (2021) doi:10.1101/2021.09.29.21264272.

24. Golubchik, T. et al. Early analysis of a potential link between viral load and the N501Y mutation in the SARS-COV-2 spike protein. medRxiv 2021.01.12.20249080 (2021) doi:10.1101/2021.01.12.20249080.

25. Funk, T. et al. Characteristics of SARS-CoV-2 variants of concern B.1.1.7, B.1.351 or P.1: data from seven EU/EEA countries, weeks 38/2020 to 10/2021. Eurosurveillance 26, (2021).

26. Greaney, A. J. et al. Mapping mutations to the SARS-CoV-2 RBD that escape binding by different classes of antibodies. Nature Communications 12, 4196 (2021).

27. Wang, P. et al. Antibody resistance of SARS-CoV-2 variants B.1.351 and B.1.1.7. Nature 593, 130–135 (2021).

28. Chen, R. E. et al. Resistance of SARS-CoV-2 variants to neutralization by monoclonal and serum-derived polyclonal antibodies. Nature Medicine 27, 717–726 (2021).

29. Liu, Z. et al. Identification of SARS-CoV-2 spike mutations that attenuate monoclonal and serum antibody neutralization. Cell Host & Microbe 29, 477-488.e4 (2021).

30. Choi, B. et al. Persistence and Evolution of SARS-CoV-2 in an Immunocompromised Host. New England Journal of Medicine 383, 2291–2293 (2020).

31. Xiao, M. et al. Multiple approaches for massively parallel sequencing of SARS-CoV-2 genomes directly from clinical samples. Genome Medicine 12, 57 (2020).

32. Charre, C. et al. Evaluation of NGS-based approaches for SARS-CoV-2 whole genome characterisation. Virus Evolution 6, (2020).

33. Addetia, A. et al. Sensitive Recovery of Complete SARS-CoV-2 Genomes from Clinical Samples by Use of Swift Biosciences’ SARS-CoV-2 Multiplex Amplicon Sequencing Panel. Journal of Clinical Microbiology 59, e02226–20 (2020).

34. Food and Drug Administration. Considerations for Design, Development, and Analytical Validation of Next Generation Sequencing (NGS) – Based In Vitro Diagnostics (IVDs) Intended to Aid in the Diagnosis of Suspected Germline Diseases. https://www.fda.gov/media/99208/download (2018).

35. Food and Drug Administration. Submitting Next Generation Sequencing Data to the Division of Antiviral Products Guidance for Industry Technical Specifications Document. hFood and Drug (2019).

36. Sadoff, J. et al. Interim Results of a Phase 1–2a Trial of Ad26.COV2.S Covid-19 Vaccine. New England Journal of Medicine 384, 1824–1835 (2021).

37. Sadoff, J. et al. Safety and Efficacy of Single-Dose Ad26.COV2.S Vaccine against Covid-19. New England Journal of Medicine 384, 2187–2201 (2021).

38. Hilt, E. E. et al. Retrospective Detection of Severe Acute Respiratory Syndrome Coronavirus 2 (SARS-CoV-2) in Symptomatic Patients Prior to Widespread Diagnostic Testing in Southern California. Clinical Infectious Diseases (2021) doi:10.1093/cid/ciab360.

39. Lin, M. J. et al. Host-pathogen dynamics in longitudinal clinical specimens from patients with COVID-19. medRxiv 2021.04.27.21256149 (2021) doi:10.1101/2021.04.27.21256149.

40. McEwen, A. E. et al. Variants of concern are overrepresented among post-vaccination breakthrough infections of SARS-CoV-2 in Washington State. medRxiv 2021.05.23.21257679 (2021) doi:10.1101/2021.05.23.21257679.

41. A., L. J. et al. Comparison of Commercially Available and Laboratory-Developed Assays for In Vitro Detection of SARS-CoV-2 in Clinical Laboratories. Journal of Clinical Microbiology 58, e00821–20 (2021).

42. Degli-Angeli, E. et al. Validation and verification of the Abbott RealTime SARS-CoV-2 assay analytical and clinical performance. Journal of Clinical Virology 129, 104474 (2020).

43. Jane, K. et al. Comparison of Real-Time PCR Assays with Fluorescent-Antibody Assays for Diagnosis of Respiratory Virus Infections in Children. Journal of Clinical Microbiology 44, 2382–2388 (2006).

44. Bruce, E. A. et al. Predicting Infectivity: Comparing Four PCR-based Assays to Detect Culturable SARS-CoV-2 in Clinical Samples. medRxiv 2021.07.14.21260544 (2021) doi:10.1101/2021.07.14.21260544.

45. Graham, N. R. et al. Kinetics and isotype assessment of antibodies targeting the spike protein receptor-binding domain of severe acute respiratory syndrome-coronavirus-2 in COVID-19 patients as a function of age, biological sex and disease severity. Clinical & Translational Immunology 9, e1189 (2020).

46. K., N. A. et al. Comparative Performance of SARS-CoV-2 Detection Assays Using Seven Different Primer-Probe Sets and One Assay Kit. Journal of Clinical Microbiology 58, e00557–20 (2021).

47. L., G. A. et al. Rapid Metagenomic Next-Generation Sequencing during an Investigation of Hospital-Acquired Human Parainfluenza Virus 3 Infections. Journal of Clinical Microbiology 55, 177–182 (2017).

48. Peddu, V. et al. Metagenomic Analysis Reveals Clinical SARS-CoV-2 Infection and Bacterial or Viral Superinfection and Colonization. Clinical Chemistry 66, 966–972 (2020).

49. Danecek, P. et al. Twelve years of SAMtools and BCFtools. GigaScience 10, p(2021).

50. R Core Team. R: A language and environment for statistical computing. R Foundation for Statistical Computing. (2014).

51. Burd, E. M. Validation of laboratory-developed molecular assays for infectious diseases. Clinical microbiology reviews 23, 550–576 (2010).

52. Chiara, M. et al. Next generation sequencing of SARS-CoV-2 genomes: challenges, applications and opportunities. Briefings in Bioinformatics (2020) doi:10.1093/bib/bbaa297.

53. Tyson, J. R. et al. Improvements to the ARTIC multiplex PCR method for SARS-CoV-2 genome sequencing using nanopore. bioRxiv 2020.09.04.283077 (2020) doi:10.1101/2020.09.04.283077.

54. Itokawa, K., Sekizuka, T., Hashino, M., Tanaka, R. & Kuroda, M. Disentangling primer interactions improves SARS-CoV-2 genome sequencing by multiplex tiling PCR. PLOS ONE 15, e0239403 (2020).

55. Addetia, A. et al. Identification of multiple large deletions in ORF7a resulting in in-frame gene fusions in clinical SARS-CoV-2 isolates. Journal of clinical virology□: the official publication of the Pan American Society for Clinical Virology 129, 104523 (2020).

