## Supplementary Figures for "Clinical performance characteristics of the Swift Normalase Amplicon Panel for sensitive recovery of SARS-CoV-2 genomes"

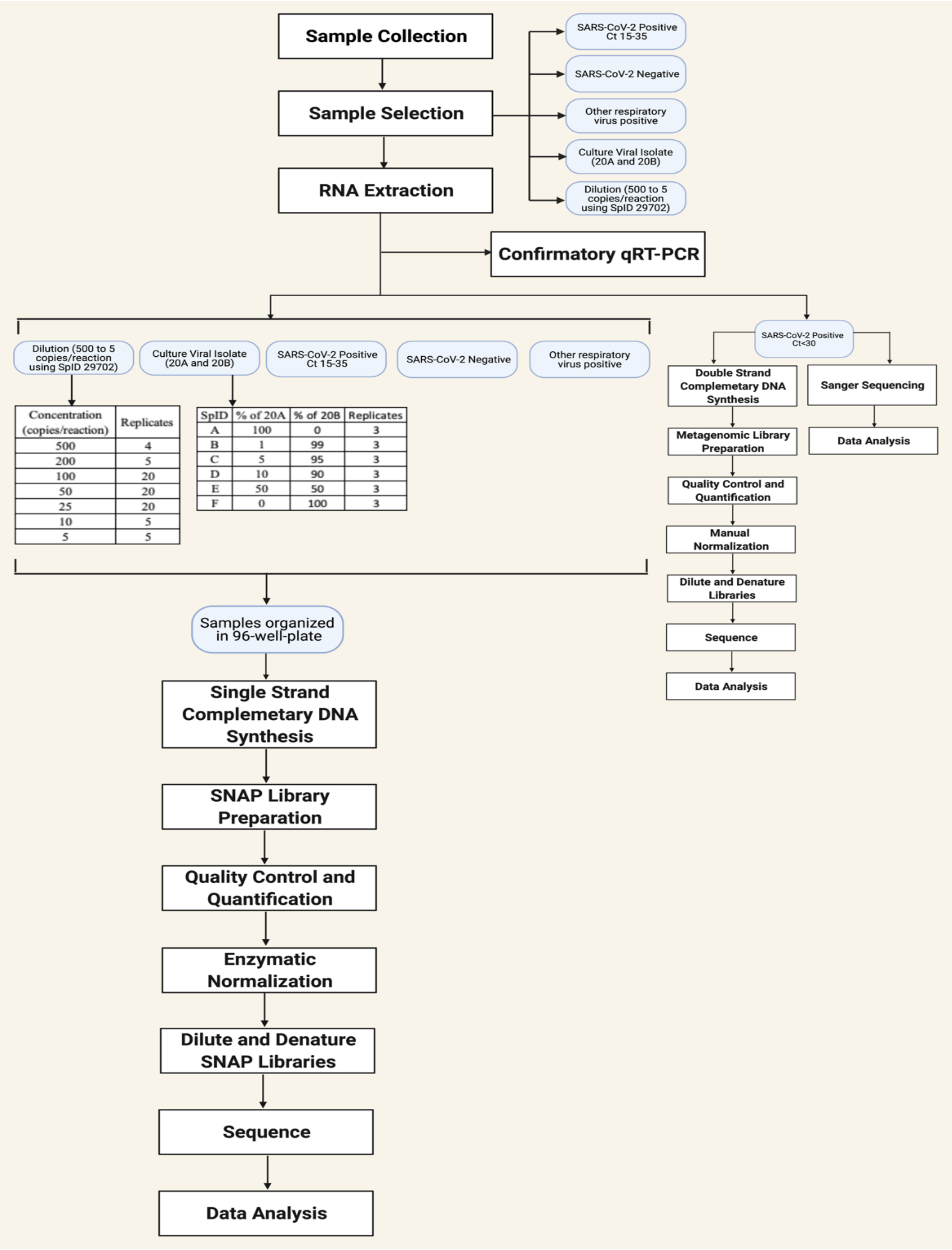


| Concentration (copies/reaction) | Replicates |
|---|---|
| 500 | 4 |
| 200 | 5 |
| 100 | 20 |
| 50 | 20 |
| 25 | 20 |
| 10 | 5 |
| 5 | 5 |

| SpID | % of 20A | % of 20B | Replicates |
|---|---|---|---|
| A | 100 | 0 | 3 |
| B | 1 | 99 | 3 |
| C | 5 | 95 | 3 |
| D | 10 | 90 | 3 |
| E | 50 | 50 | 3 |
| F | 0 | 100 | 3 |

Supplementary Figure 1.

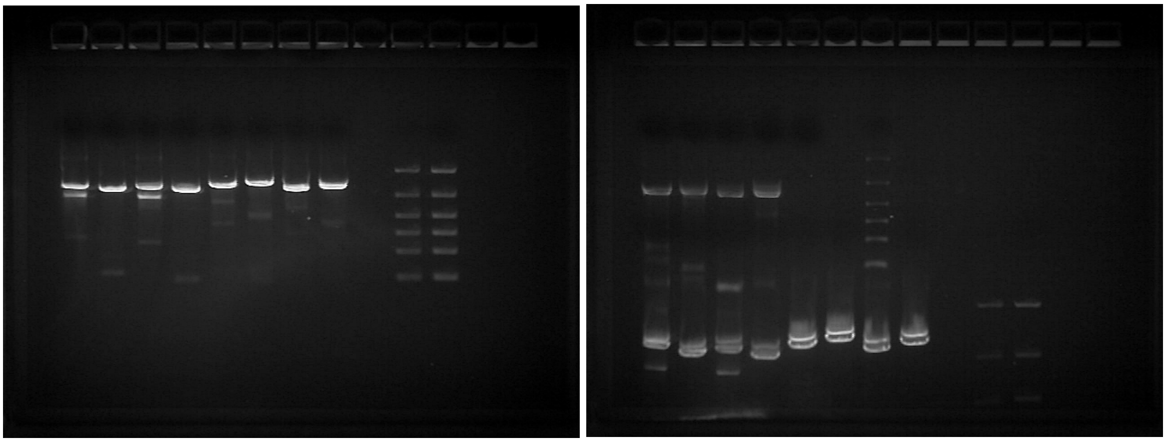

**Supplementary Figure 2.** Gels showing Sanger amplicons for one representative sample (SpID 26865). Lanes from L to R show: (gel 1) Amplicon 1 F1a/R1a, F1a/R1b, F1b/R1a, F1b/R1b; Amplicon 2 F2a/R2a, F2a/R2b, F2b/R2a, F2b/R2b; blank lane; 3µL of 100-3000 bp ladder; 5µL ladder; (gel 2) Amplicon 3 F3a/R3a, F3a/R3b, F3b/R3a, F3b/R3b, two blank lanes; 3µL ladder. Note that the Amplicon 1 and Amplicon 2 samples from gel 1 are visible at the bottom of gel 2 because samples were run consecutively on the same gel. Most sequencing was performed on amplicons generated using primers F1a/R1b, F2a/R2a, and F3b/R3b.

University of Washington
Laboratory Medicine

Department of Laboratory Medicine · UW Medicine · Box 357110 · 1959 NE Pacific Street · Seattle, WA 98195-7110 · Tel: 1-800-713-5198

| | |
|---|---|
| Collected [redacted] | Patient [redacted] |
| Received [redacted] | DOB [redacted] |
| Client MRN [redacted] | UWMC MRN [redacted] |
| Client Acc. [redacted] | UWMC Acc. [redacted] |
| Reported [redacted] | Ordered by [redacted] |

### SARS-CoV-2 Whole Genome Sequencing Assay

#### Strain typing and resistance testing

**NGS Variant Analysis**
20J (Gamma, V3)/P.1

Interpretation
The sequence variants identified place the isolate in the 20J (Gamma, V3) clade, specifically the P.1 lineage. This strain is considered a Variant of Concern by the SARS-CoV-2 Interagency Group.

**SARS-CoV-2 NGS Spike Gene Mutations**
L18F, T20N, P26S, D138Y, R190S, K417T, E484K, N501Y, D614G, H655Y, T1027I, V1176F

Interpretation
L18F, T20N, P26S, D138Y, R190S, K417T, E484K, N501Y, D614G, H655Y, T1027I and V1176F amino acid variants detected in the Spike gene. N501Y is associated with increased human ACE2 binding and increased transmissibility. E484K is associated with antibody escape.

**Reference**
Reference sequence used for all analyses is NC_045512.2 from NCBI Genbank and the name of the reference is WuHan_Hu1.

This test was developed and its performance characteristics determined by the University of Washington Department of Laboratory Medicine and Pathology. It has not been cleared or approved by the US Food and Drug Administration. This laboratory is certified under the Clinical Laboratory Improvement Amendments (CLIA) as qualified to perform high complexity clinical laboratory testing. This test is used for clinical purposes. It should not be regarded as investigational or for research.

Testing performed by the UW Medicine Virology Lab located at 1616 Eastlake Ave. E, Ste 320, Seattle, WA 98102, Keith Jerome, MD, PhD, Lab Director

End of report



**Supplementary Figure 3.** Sample report for clinical test orders

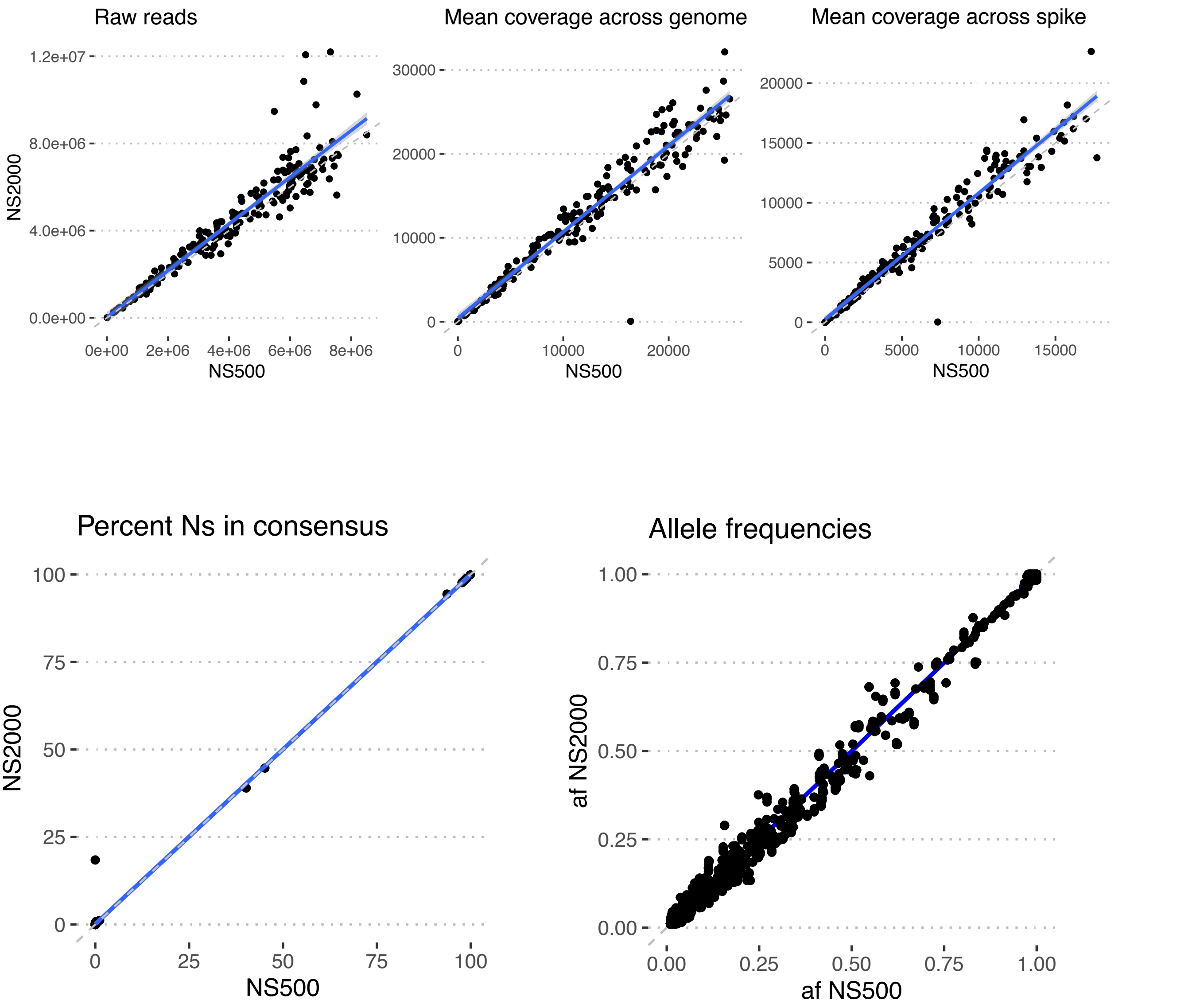


**Supplementary Figure 4.** Instrument validation: identical libraries (n = 181) were sequenced in parallel on the Illumina NextSeq 500 and an Illumina NextSeq 2000 (VH00441).

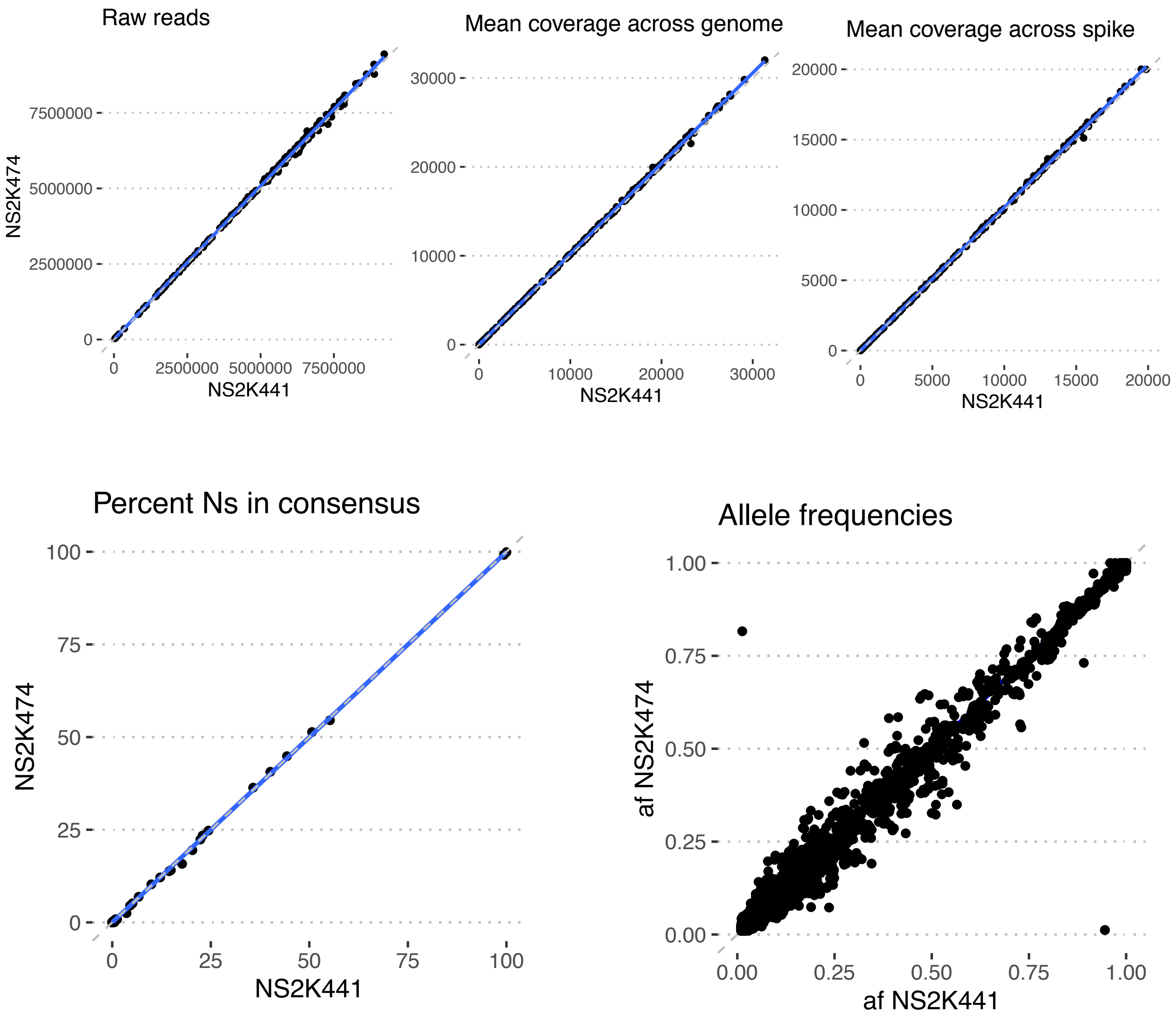


**Supplementary Figure 5.** Instrument validation: identical libraries (n = 185) were sequenced in parallel on two different Illumina NextSeq 2000 instruments (VH00441/NS2K441 vs VH00474/NS2K474)

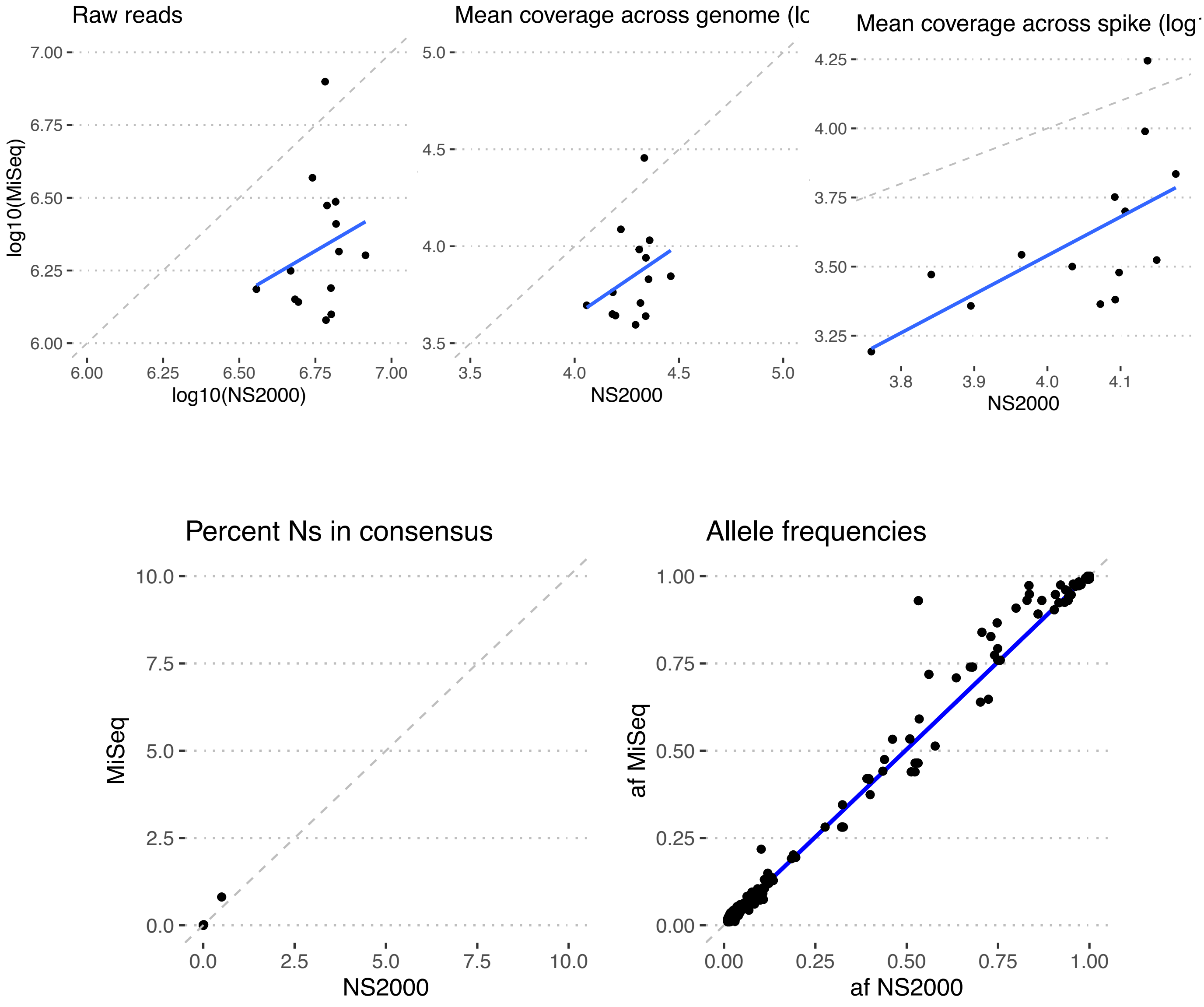


**Supplementary Figure 6.** Instrument validation: identical libraries (n = 12) were sequenced in parallel on an Illumina NextSeq 2000 and an Illumina MiSeq.
